# Effect of the 2022 COVID-19 booster vaccination campaign in 50 year olds in England: regression discontinuity analysis in OpenSAFELY

**DOI:** 10.1101/2023.09.07.23295194

**Authors:** Andrea L Schaffer, William J Hulme, Elsie Horne, Edward P K Parker, Venexia Walker, Catherine Stables, Amir Mehrkar, Seb CJ Bacon, Chris Bates, Ben Goldacre, Alex J Walker, The OpenSAFELY Collaborative, Miguel A Hernán, Jonathan A C Sterne

## Abstract

**Background:** SARS-CoV-2 vaccines are highly effective in preventing severe COVID-19 but require boosting to maintain protection. Changes to circulating variants and prevalent natural immunity may impact on real-world effectiveness of boosters in different time periods and in different populations.

**Methods:** With NHS England approval, we used linked routine clinical data from >24 million patients to evaluate the effectiveness of the 2022 combined COVID-19 autumn booster and influenza vaccine campaign in non-clinically vulnerable 50-year-olds in England using a regression discontinuity design. Our primary outcome was a composite of 6-week COVID-19 emergency attendance, COVID-19 unplanned hospitalisation, or death. The secondary outcomes were: respiratory hospitalisations or death; any unplanned hospitalisation; and any death.

**Results:** Our study included 1,917,375 people aged 45-54 years with no evidence of being in a high-risk group prioritised for vaccination. By 26 November 2022, booster vaccine coverage was 11.1% at age 49.75 years increasing to 39.7% at age 50.25 years. The estimated effect of the campaign on the risk of the primary outcome in 50-year-olds during weeks 7-12 after the campaign start was -0.4 per 100,000 (95% CI -7.8, 7.1). For the secondary outcomes the estimated effects were: -0.6 per 100,000 (95%CI -13.5, 12.3) for respiratory outcomes; 5.0 per 100,000 (95%CI -40.7, 50.8) for unplanned hospitalisations; and 3.0 per 100,000 (95%CI -2.7, 8.6) for any death. The results were similar when using different follow-up start dates, different bandwidths, or when estimating the effect of vaccination (rather than the campaign).

**Conclusion:** This study found little evidence that the autumn 2022 vaccination campaign in England was associated with a reduction in severe COVID-19-related outcomes among non-clinically vulnerable 50-year-olds. Possible explanations include the low risk of severe outcomes due to substantial pre-existing vaccine- and infection-induced immunity. Modest booster coverage reduced the precision with which we could estimate effectiveness. The booster campaign may have had effects beyond those estimated, including reducing virus transmission and incidence of mild or moderate COVID-19.

## Background

The SARS-CoV-2 vaccines are highly effective at preventing severe COVID-19 outcomes, including hospitalisation and mortality.(1,2) However, their benefits wane over time(3) and so booster vaccinations are needed to sustain protection. A first booster vaccination reduces the incidence of severe COVID-19, but that protection also wanes and may be reduced against new variants.(4–9) High prevalence of immunity resulting from prior SARS-CoV-2 infection may impact on booster vaccine effectiveness.

Following the primary course of two doses, the UK offered first COVID-19 booster vaccinations from autumn 2021 to high-risk individuals and people 50 years and older. A spring 2022 booster was subsequently offered to people 75 years or older and those considered clinically vulnerable. An autumn booster was available from September 2022, initially targeting people considered at high risk, such as those aged over 65 years, care home residents and their staff, and immunosuppressed people.(10,11) On 15 October 2022, people aged 50-64 years who were not considered high risk became eligible for booster vaccination.(12) This coincided with the 2022/23 rollout of influenza vaccination, which was available from the same date to people who would turn 50 by March 2023.(12)

In this study, we estimated the effectiveness of the combined 2022 autumn COVID-19 booster vaccination campaign, coinciding with the influenza vaccination campaign, in reducing COVID-19 outcomes among non-clinically vulnerable people aged 50 years in England using a regression discontinuity design and the OpenSAFELY-TPP database.

## Methods

### Study objective and population

Our primary objective was to estimate the average effect of the combined COVID-19 booster and influenza vaccination campaign on severe COVID-19 outcomes in non-clinically vulnerable people aged 50 years in England who had previously received at least two COVID-19 vaccinations. This population became eligible for the COVID-19 booster and influenza vaccine on 15 October 2022. Our secondary objective was to estimate the effect of the booster vaccine itself among compliers; that is, people who take up the vaccine only when eligible.

We included all adults aged 45-54 years during the study period who were registered at one GP practice for at least 90 days prior to 3 September 2022, and had complete information on age and sex. People considered high risk or clinically vulnerable, who were eligible for booster vaccination earlier in the year were excluded.(10) This included people who were(13): identified as a health or social care workers at time of vaccination; resident in a care or nursing home; or part of any other clinically vulnerable group, specifically with: chronic respiratory disease, chronic heart/vascular disease, chronic kidney disease, chronic liver disease, chronic neurological disease, diabetes, immunosuppression, asplenia, morbid obesity, or severe mental illness; or with evidence of having received a third primary dose of the COVID-19 vaccine which may be a marker of immunosuppression.

We also excluded people who were otherwise ineligible for the booster or unlikely to be vaccinated, including people who: received another COVID-19 vaccine within 90 days prior to 15 October 2022; did not receive the first two primary doses of the COVID-19 vaccine; were housebound; or were receiving end of life care. **Supplementary Table 1** lists the exclusion criteria and their definitions. Clinically vulnerable individuals were identified using primary care data using the same approach as described previously.(3,14)

### Data Source

All data were linked, stored and analysed securely within the OpenSAFELY platform: https://opensafely.org/. With the approval of NHS England, primary care records managed by the GP software provider TPP were linked, using NHS numbers, to accident and emergency attendance (A&E) and in-patient hospital spell records via NHS Digital’s Hospital Episode Statistics (HES), and national death registry records from the Office for National Statistics (ONS). COVID-19 vaccination history is available in the GP record directly via the National Immunisation Management System (NIMS). The dataset analysed within OpenSAFELY is based on 24 million people currently registered with GP surgeries using TPP SystmOne software.

### Study Measures

#### Outcomes

The primary outcome was a composite of COVID-19-related unplanned hospitalisation, COVID-19-related accident and emergency attendance, or any death from 6-weeks after start of the vaccination campaign. For unplanned hospitalisations, the COVID-19 diagnosis could be either an underlying or contributing cause in any position. Only unplanned admissions were included as these are more likely to be due to incident COVID-19 disease. We did not include a positive SARS-CoV-2 test as an outcome because free testing in England ended in April 2022.

Due to potential misdiagnosis of COVID-19 related outcomes, we also included a composite outcome of respiratory unplanned admission or death. We also examined any unplanned hospitalisations and any death. The definition and codes used to identify outcomes are in **Supplementary Table 2**.

#### Exposure

The exposure was booster eligibility, defined as being 50 years or older on the start of follow-up, for vaccination in the autumn 2022 booster campaign. Age was categorised in 3-month intervals. As only month of birth is available in OpenSAFELY, date of birth was set to the 15th of the month. We defined receipt of the autumn booster as a record of a third or fourth COVID-19 vaccination on or after 5 September 2022 (the date autumn boosters first became available), as some people may have been vaccinated prior to eligibility. We constructed cumulative incidence curves of booster coverage to display how separation above/below the threshold changed over the study period.

### Study design

We used regression discontinuity, a study design that takes advantage of the threshold of being aged 50 years or older for booster vaccination eligibility and estimates the effectiveness of booster vaccination at this threshold. Threshold-based eligibility mimics randomisation, as the distribution of confounding variables among people just above and below the threshold is expected to be similar.(15) Regression discontinuity has previously been used to estimate the effectiveness of vaccines and vaccination campaigns, such as the first COVID-19 vaccine dose on COVID-19 mortality in England(16) and influenza vaccination in England and Wales.(17)

The population-level effect of booster vaccination is likely to evolve over time, as the proportion of people vaccinated increases, and the prevalence of SARS-CoV-2 infection changes. We therefore included multiple index dates for the start of follow-up as supplementary analyses: 3 September 2022 (before the start of the campaign; negative control); 15 October 2022 (start of the campaign; negative control); and each day between 26 November (6 weeks after the start of the campaign) and 9 December 2022. Follow-up was for 6 weeks after each index date. For each index date, we excluded people who had died or deregistered before that date.

#### Assumptions

Key assumptions of regression discontinuity are “continuity” (the risk of the outcome is expected to change smoothly at the threshold in absence of the intervention) and “exchangeability” (similar distribution of confounders just below and above the threshold).(15) To test the latter assumption, we plotted the distribution of the following characteristics by age in 3 month intervals: sex (male, female); deprivation, measured by the English Index of Multiple Deprivation (IMD), grouped by quintile of national rank; ethnicity (White, Mixed, Asian/Asian British, Black/Black British, Other, Unknown); practice region (East, East Midlands, London, North East, North West, South East, South West, Yorkshire and The Humber); number of previous COVID-19 vaccine doses. We also quantified receipt of influenza vaccine in the 2022/23 season from July 2022 onwards to examine its discontinuity at threshold. However, ascertainment of influenza vaccination was incomplete, because vaccination delivered by pharmacists outside of general practice is not routinely recorded in electronic health records.

We used both “sharp” and “fuzzy” regression discontinuity designs, which address different research questions.(18,19) The sharp design estimates the effect of the vaccination campaign (rather than receipt of vaccination itself) at the threshold age. As both the second booster and the influenza vaccine became available on the same date at the same age threshold, and thus there is an expected discontinuity in receipt of flu vaccination, this represents the effect of the combined vaccination campaigns.

The fuzzy design estimates the effectiveness of receiving booster vaccination during autumn 2022: those boosted commonly received influenza vaccination as well. Fuzzy regression discontinuity uses vaccine eligibility (being age 50 years or older) as an instrumental variable and estimates the “local average treatment effect” (LATE) or “complier average causal effect” (CACE) at the threshold.(15) This is the effect of vaccination among “compliers” - the subset of the population who are vaccinated only when eligible. The analysis assumes no “defiers” - people who are vaccinated only when ineligible. This assumption is also known as monotonicity. The monotonicity assumption is untestable, but likely to hold in our study. Other assumptions are that the instrumental variable is associated with the outcome only through vaccination at the threshold, that there are no common causes of eligibility and the outcome.(20) These assumptions are also plausible for our study.

### Statistical methods

Outcomes were expressed as 6-week risks per 100,000 population. To prevent disclosure, all event counts presented were rounded to the nearest 5 and risks calculated from rounded counts. However, unrounded counts and risks were used in regression modelling. For each index date and follow-up period, only the first event for each individual was counted. The primary bandwidth was 5 years (20 data points representing three-month age intervals) either side of the threshold.

To estimate the effect of being eligible for the COVID-19 autumn booster/influenza vaccine among people aged 50 years (sharp regression discontinuity), we estimated the discontinuity in the risk of each outcome at the threshold by fitting a regression model with age in 3-month intervals (continuous), a binary variable representing the vaccine age threshold (50+ years), and an interaction between the two, allowing the slope of the change in the probability of the outcome by age to vary above and below the threshold. Age was centred, so that “0” represented the threshold.

We next used an instrumental variable approach to estimate the effect of vaccination in people aged 50 who were vaccinated (fuzzy regression discontinuity) with the instrument being 50 years or older on the index date (eligible for vaccination). We used the two-stage least squares estimator. In the first stage, booster coverage before the index date is predicted based on age. The second stage uses the predicted values from the first stage to predict the outcome.(15)

#### Sensitivity analyses

For the sharp regression discontinuity analysis, we conducted two sensitivity analyses: first, we excluded people born in the index month due to the imprecision in recorded birth date. Second, given the potential for bias with wide bandwidths, we repeated the analysis using progressively smaller bandwidths (4 years, 3 years, 2 years, 1 year). For the fuzzy regression discontinuity analysis, we repeated the analysis including influenza vaccination prior to the index date in the model, and also using different bandwidth periods.

### Software and Reproducibility

Data management was performed using Python 3.8, with analysis carried out using R. Code for data management and analysis as well as codelists archived online https://github.com/opensafely/vax-fourth-dose-RD.

### Patient and Public Involvement

We have developed a publicly available website https://opensafely.org/ through which we invite any patient or member of the public to contact us regarding this study or the broader OpenSAFELY project.

### Ethics approval

This study was approved by the Health Research Authority (Research Ethics Committee reference 20/LO/0651) and by the London School of Hygiene and Tropical Medicine Ethics Board (reference 21863).

## Results

We identified 3,254,000 people aged 45-54 years on 3 September 2022. Of these, 1,336,625 (41.1%) were excluded (**Table 1**). The most common reasons for exclusion were not having received a second primary COVID-19 dose (521,165, 16.0%) or being in a clinically vulnerable group (762,650, 23.4%), most frequently due to diabetes (229,905, 7.1%), chronic heart disease (160,165, 4.9%) or severe obesity (143,720, 4.4%).

**Table 1.** All people 45-54 years on 3 September 2022 and reasons for exclusion from the study. People could have multiple exclusion criteria and so may appear in multiple categories. All counts rounded to nearest 5.

|  | N (%)^ |
| --- | --- |
| **Total prior to exclusions** | 3,254,000 (100.0) |
| **Any exclusion** | 1,336,625 (41.1) |
| **Included in final cohort** | 1,917,375 (58.9) |
| **Prioritised for vaccination** |  |
| Living in care home | 7595 (0.2) |
| Health and social care worker | 149,070 (4.6) |
| Clinically vulnerable | 762,650 (23.4) |
| Immunosuppressed | 99,370 (3.1) |
| Chronic kidney disease | 29,760 (0.9) |
| Chronic respiratory disease | 60,345 (1.9) |
| Asthma | 14,440 (0.4) |
| Diabetes | 229,905 (7.1) |
| Asplenia | 19,130 (0.6) |
| Chronic liver disease | 92,525 (2.8) |
| Chronic heart disease | 160,165 (4.9) |
| Severe mental illness | 41,680 (1.3) |
| Severe obesity | 143,720 (4.4) |
| **Other reasons for exclusion** |  |
| Receiving end of life care | 5055 (0.2) |
| Housebound | 6440 (0.2) |
| Received COVID-19 vaccine within 90 days prior to campaign start | 20,655 (0.6) |
| Received 3<sup>rd</sup> COVID-19 vaccine prior to wider availability* | 5745 (0.2) |
| Received 4<sup>th</sup> COVID-19 vaccine prior to wider availability* | 59,660 (1.8) |
| Did not receive second primary COVID-19 vaccine dose | 521,165 (16.0) |

^n = 3,254,000 used as the denominator to calculate percentages
\*As a proxy for immunosuppressed individuals who were eligible for a third primary dose

Among 1,917,375 people included in analyses, 465,730 (51.4%) of people 45-49 years and 516,310 (51.1%) of people 50-54 years were male and 661,145 (73.0%) and 776,560 (76.8%) were of White ethnicity, respectively (**Table 2**). Most (743,485 [82.1%] and 885,095 [87.5%]) had received three previous COVID-19 vaccine doses. The median time since the previous dose at the start of the campaign (15 October) was 304 days (interquartile range [IQR], 297-321 days) and 310 days (IQR, 301-328), respectively.

**Table 2.** Demographic characteristics of people 45-54 years in final cohort on 3 September 2022. All counts rounded to nearest 5.

|  | 45-49 years | 50-54 years |
| --- | --- | --- |
|  | N (%) | N (%) |
| **Total population** | 906,040 (100.0) | 1,011,330 (100.0) |
| **Number of previous SARS-CoV-2 vaccine doses** |  |  |
| 2 doses | 162,560 (17.9) | 126,235 (12.5) |
| 3 doses | 743,485 (82.1) | 885,095 (87.5) |
| **Sex** |  |  |
| Male | 465,730 (51.4) | 516,310 (51.1) |
| Female | 440,320 (48.6) | 495,020 (48.9) |
| **Index of Multiple Deprivation (IMD) quintile** |  |  |
| 1 (most deprived) | 141,430 (15.6) | 148,365 (14.7) |
| 2 | 161,920 (17.9) | 177,060 (17.5) |
| 3 | 188,555 (20.8) | 215,485 (21.3) |
| 4 | 193,865 (21.4) | 223,280 (22.1) |
| 5 (least deprived) | 195,295 (21.6) | 223,155 (22.1) |
| Missing | 24,975 (2.8) | 23,985 (2.4) |
| **Ethnicity** |  |  |
| White | 661,145 (73.0) | 776,560 (76.8) |
| Asian or Asian British | 75,345 (8.3) | 55,080 (5.4) |
| Black | 21,365 (2.4) | 18,760 (1.9) |
| Mixed | 10,335 (1.1) | 8395 (0.8) |
| Other | 19,970 (2.2) | 15,350 (1.5) |
| Unknown | 117,875 (13.0) | 137,175 (13.6) |
| **Practice region** |  |  |
| East | 219,695 (24.2) | 237,570 (23.5) |
| East Midlands | 156,335 (17.3) | 179,650 (17.8) |
| London | 62,325 (6.9) | 55,830 (5.5) |
| North East | 38,995 (4.3) | 46,130 (4.6) |
| North West | 75,125 (8.3) | 89,675 (8.9) |
| South East | 60,165 (6.6) | 67,150 (6.6) |
| South West | 131,115 (14.5) | 152,455 (15.1) |
| West Midlands | 32,595 (3.6) | 35,730 (3.5) |
| Yorkshire and The Humber | 125,795 (14.0) | 144,355 (14.3) |
| Missing | 2900 (0.3) | 2795 (0.3) |

### Vaccination coverage

A small number of people in our study population received the booster vaccine prior to its wider availability on 15 October 2022 (**Figure 1A**). Coverage increased rapidly thereafter and started to plateau by late November (**Figure 1A**). Six weeks after the wider availability (26 November), booster coverage ranged from 6.1% of people 45.00 years to 51.8% of people aged 54.75 years (**Figure 1B**). A large discontinuity was observed at the threshold, with the proportion receiving booster vaccination increasing from 11.1% of people aged 49.75 years to 31.1% and 39.7% of people aged 50.00 and 50.25 years respectively.

**Figure 1.**
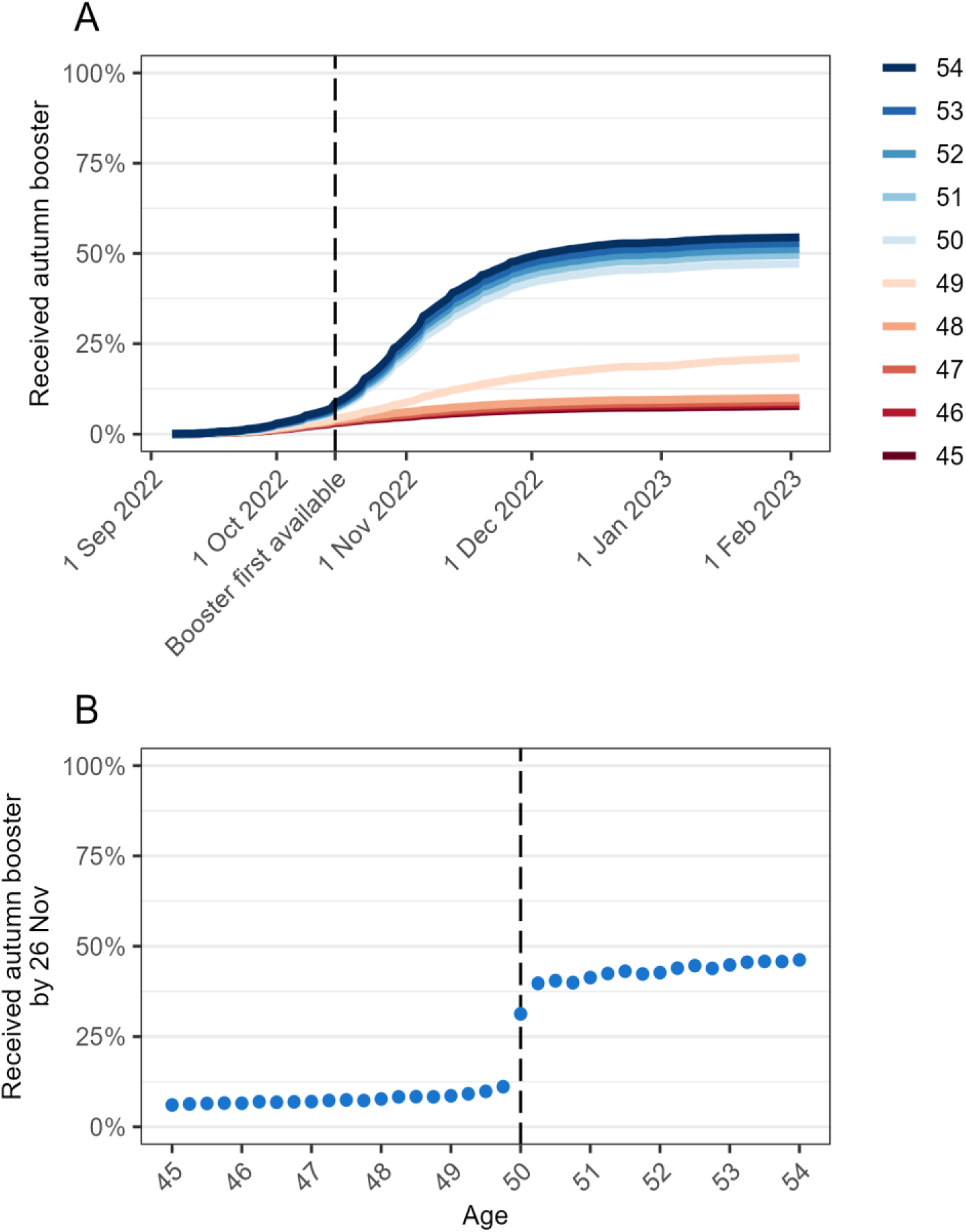
Coverage (%) of COVID-19 autumn booster by age. A) Cumulative coverage based on age in years at 3 September 2022. The booster vaccination became available to non-clinically vulnerable people aged 50-64 years on 15 October 2022. B) Coverage at 26 November 2022 by age in 3-month intervals.

No discontinuity was observed for any of the demographic variables (sex, IMD quintile, ethnicity, region) (**Supplementary Figures 1-4**) or number of prior COVID-19 vaccine doses (**Supplementary Figure 5**). However, we observed a discontinuity in recorded receipt of the 2022/23 influenza vaccine, increasing from 9.7% of people aged 49.25 years to 25.5% of people aged 50.00 years (**Supplementary Figure 6**). Recorded receipt of influenza vaccine was much more common among people who received COVID-19 booster vaccination. In people aged 45-49 years, 36,515 (53.2%) of those receiving booster vaccination also received influenza vaccination, compared with 46,820 (5.6%) of those who did not receive booster vaccination. Corresponding figures in people aged 50-54 years were 277,745 (64.7%) and 68,340 (11.8%).

### Regression discontinuity analysis

Overall, COVID-19 related outcomes (unplanned admission, A&E attendance, or death) were rare; the risks within 6 weeks of 26 November 2022 were 12.7 and 14.4 per 100,000 for people aged 45-49 and 50-54 years respectively (**Table 3**). During the same time period, the 6-week risks of respiratory unplanned admission or death were 52.0 and 53.5 per 100,000 respectively, any unplanned admission 410.5 and 447.0 per 100,000 respectively, and any death 6.1 and 13.4 per 100,000 respectively.

**Table 3.** Number of events and risks of primary and secondary outcomes with different index dates by age. All counts rounded to nearest 5.

|  | Before start of campaign<br>(3 Sep-14 Oct 2022) |  | At start of campaign<br>(15 Oct-25 Nov 2022) |  | 6 weeks after start of campaign<br>(26 Nov 2022-6 Jan 2023) |  |
| --- | --- | --- | --- | --- | --- | --- |
|  | n | Risk per 100,000 | n | Risk per 100,000 | n | Risk per 100,000 |
| **No. people** |  |  |  |  |  |  |
| 45-49 years | 906,050 | -- | 904,900 | -- | 903,680 | -- |
| 50-54 years | 1,011,330 | -- | 1,010,735 | -- | 1,010,150 | -- |
| **COVID-19 unplanned admission,**<br>**A&E attendance, or death** |  |  |  |  |  |  |
| 45-49 years | 105 | 11.6 | 110 | 12.2 | 115 | 12.7 |
| 50-54 years | 155 | 15.3 | 105 | 10.4 | 145 | 14.4 |
| **Respiratory unplanned admission**<br>**or death** |  |  |  |  |  |  |
| 45-49 years | 190 | 21.0 | 275 | 30.4 | 470 | 52.0 |
| 50-54 years | 215 | 21.3 | 300 | 29.7 | 540 | 53.5 |
| **Any unplanned admission** |  |  |  |  |  |  |
| 45-49 years | 3920 | 432.7 | 4105 | 453.6 | 3710 | 410.5 |
| 50-54 years | 4490 | 444.0 | 4800 | 474.9 | 4515 | 447.0 |
| **Any death** |  |  |  |  |  |  |
| 45-49 years | 50 | 5.5 | 55 | 6.1 | 55 | 6.1 |
| 50-54 years | 90 | 8.9 | 105 | 10.4 | 135 | 13.4 |

A&E = accident and emergency

By age, during the six weeks from 26 November 2022 the risk of the COVID-19 composite outcome was relatively constant between ages 45 and 55 years, with a slight negative slope before age 50 and a slight positive slope thereafter (**Figure 2**). The estimated effect of the booster vaccination campaign in 50-year-olds on the 6-week risk was -0.4 per 100,000 (95% CI -7.8 to 7.1) (**Supplementary Table 3**). Similarly, for the respiratory composite outcome the estimated effect on 6-week risk -0.6 per 100,000 (95% CI -13.5 to 12.3) and for any unplanned admission 5.0 per 100,000 (95% CI -40.7 to 50.8). For any death the effect was 3.0 per 100,000 (95% CI -2.7 to 8.6). **Figure 3** shows corresponding estimates of the effect of the booster campaign for each index date and each outcome for people at the threshold: the estimates for index date 26 November correspond to those shown in **Figure 2**. Results were similar using different index dates for all outcomes (**Supplementary Table 3**). These results also were little changed after excluding people born in the index month (**Supplementary Table 4**). The results were also robust to different bandwidths, but as expected the confidence intervals were wider for shorter bandwidths (**Supplementary Table 5**).

**Figure 2.**
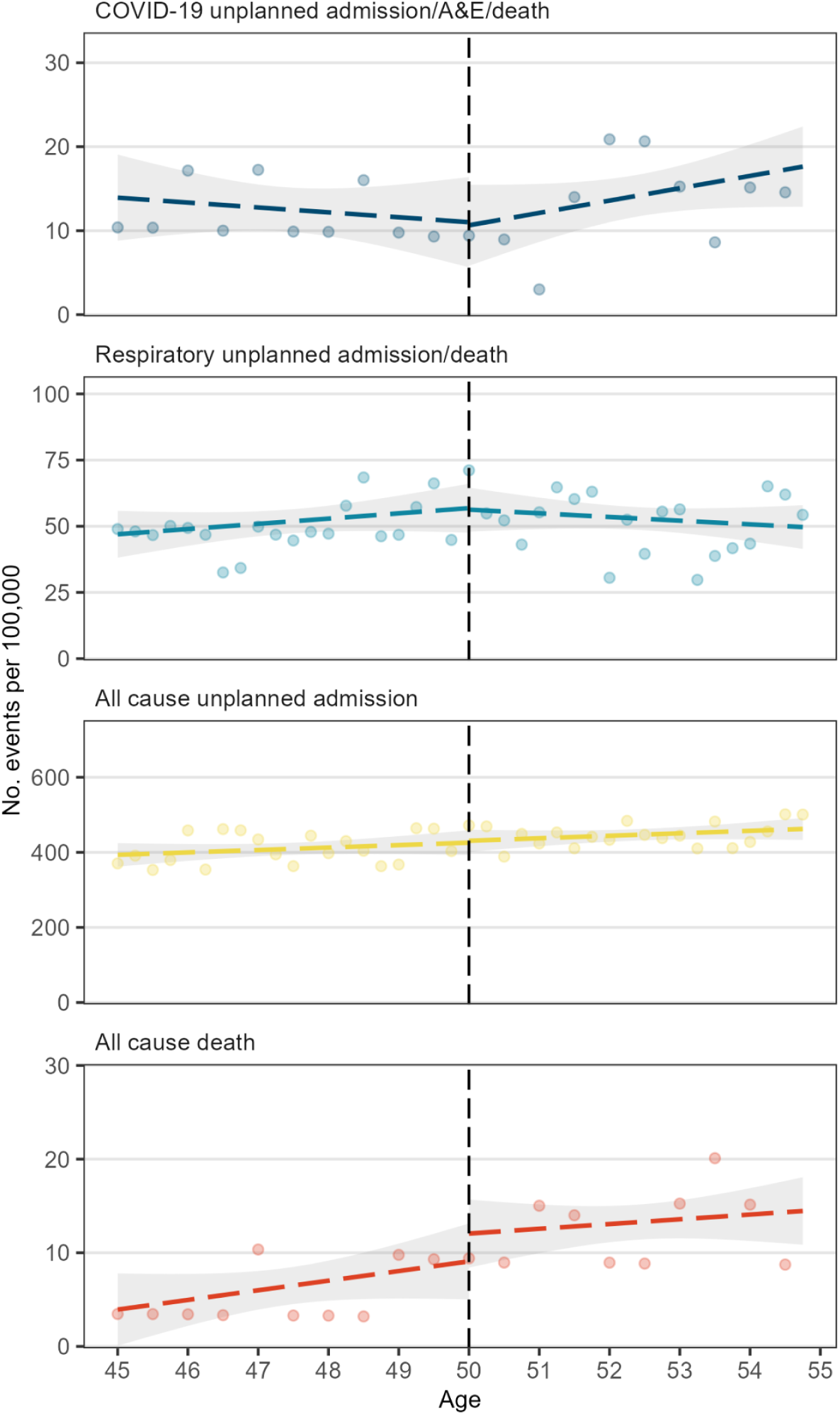
Predicted values (dashed line) and 95% confidence interval (shaded area) from sharp regression discontinuity analysis of 6-week outcomes by age in 3-month intervals with 26 November as the index date. The dots are the observed values; to prevent disclosure, rates are calculated from rounded counts and for the COVID-19 composite outcome and any death outcome, observed values for 6-month intervals are presented instead of 3 months for visualisation only.

**Figure 3.**
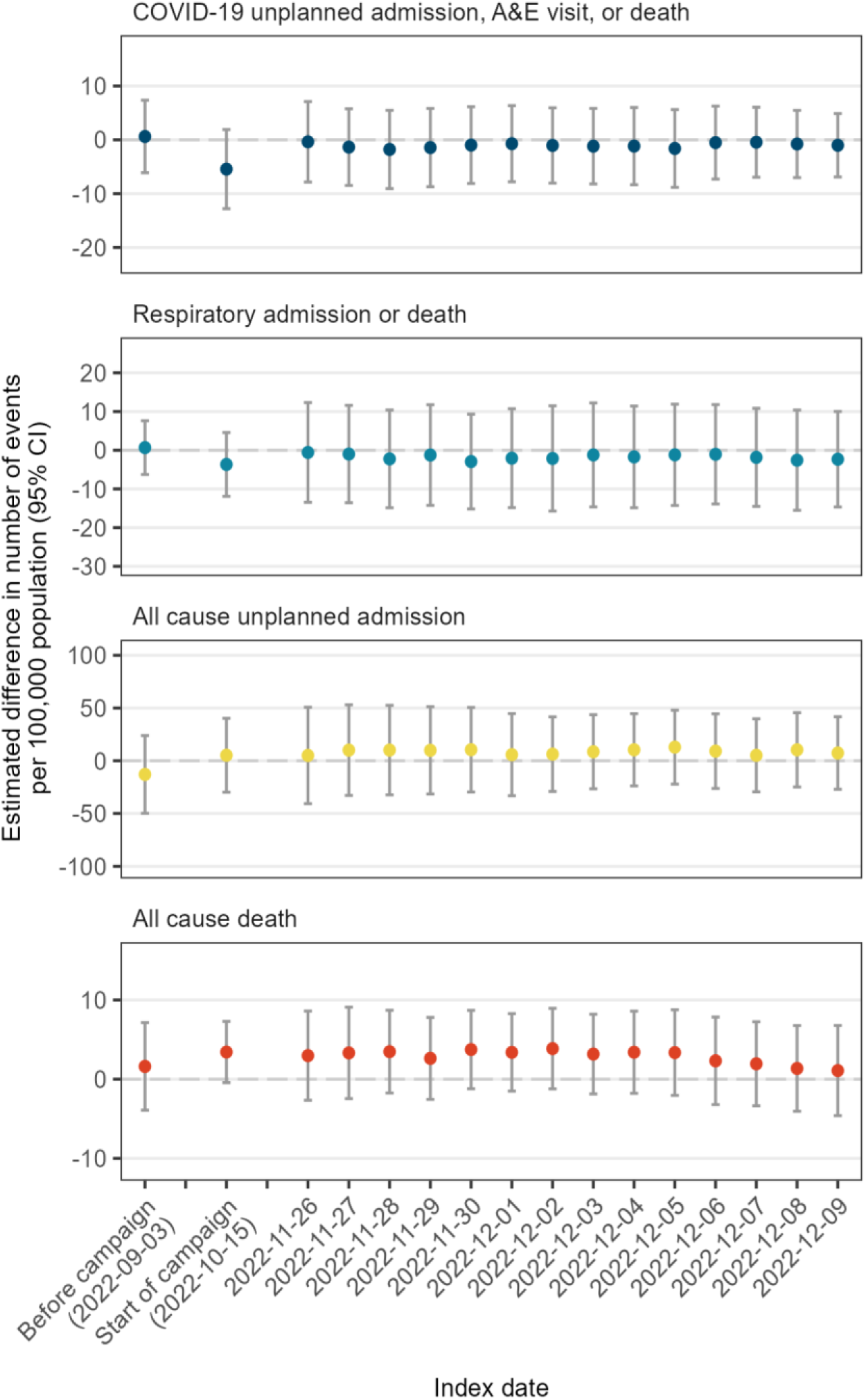
Estimated difference in 6-week outcome risk per 100,000 among people aged 50 years using different index dates. A positive estimate indicates a higher risk among people eligible to receive the booster vaccination compared with those ineligible. Whiskers represent 95% confidence intervals.

The instrumental variable analysis (fuzzy regression discontinuity) estimates the effectiveness of the booster vaccine in compliers at the threshold. Based on the first stage of the instrumental variable analysis, the estimated proportion of compliers among the population at the threshold is 28%. This analysis found a difference of 2.1 per 100,000 compliers for the COVID-19 composite outcome (95%CI -11.3 to 15.4) (**Supplementary Table 6**). This is expected given the absence of evidence for a population-level effect of the booster vaccination campaign. Controlling for prior receipt of influenza vaccination did not change the conclusions of the instrumental variable analyses, although confidence intervals became wider (**Supplementary Table 7**). Similarly, using different bandwidths did not change the findings (**Supplementary Table 8**).

## Discussion

### Summary

In this study of over 1.3 million people aged 45-54 years in England, there was a low risk of severe COVID-19 outcomes among non-clinically vulnerable people aged 45-54 years who had received at least two COVID-19 vaccine doses, and only moderate coverage of booster vaccination in people aged ≥50 years in autumn/winter 2022/23. Using a regression discontinuity design, we found little evidence of a population effect of the combined COVID-19 booster and influenza vaccination campaign on severe COVID-19 related events, unplanned respiratory or all-cause admissions or death in people aged 50 years. However, confidence intervals were wide and do not exclude a small effect.

### Strengths and weaknesses

The OpenSAFELY-TPP database covers approximately 40% of registered English primary care patients(21) enabling analysis of a large cohort of people aged within five years of the booster vaccination age eligibility threshold. These data include reliable ascertainment of COVID-19 vaccination status, COVID-19-related outcomes, and other clinical characteristics.

Establishing causality using observational data is challenging.(22) Unlike many observational study designs, regression discontinuity analyses are unlikely to be impacted by unmeasured confounding. Vaccination campaigns are well-suited to regression discontinuity designs, as eligibility is often based on a threshold, usually age. There is a reasonable assumption of “exchangeability” of people on either side of this threshold. We checked this assumption and did not find discontinuities for several potentially confounding variables (e.g. deprivation), a notable exception being receipt of influenza vaccination which was subject to the same age eligibility on the same date. Thus, the results should be considered to estimate the combined effect of the COVID-19 booster and influenza vaccination campaigns at age 50 years as we cannot disentangle the effects of the two vaccines.

Our analysis also does not account for non-independence within and across exposure groups, arising from transmissibility of SARS-CoV-2. People below age 50 may indirectly benefit from booster vaccination amongst their slightly older peers due to reduced transmission rates, resulting in lower estimates of effectiveness in the sharp discontinuity design.

### Findings in context

Various studies worldwide have demonstrated the effectiveness of a second booster against SARS-CoV-2 infection or severe COVID-19 outcomes when the Omicron variant predominates, with many identifying rapid waning.(23–25) However, the majority of studies focussed on older populations (60+ years), or do not report age-specific effectiveness estimates. Therefore, there are few studies with which to compare our findings. One previous study evaluated the 2022 autumn booster in England using a test-negative design: this found that the effectiveness of three or more COVD-19 vaccine doses against COVID-19 hospitalisation in people 18-64 years was 61.5% compared with unvaccinated people in the first two weeks post-vaccination, falling to 38.9% by 6 months.(26) Additionally, the incremental effectiveness of the booster against hospitalisation among people 50+ years who had previously received at least two doses was estimated to be 43.7% and 47.5% at 5-9 weeks post-vaccination for the Pfizer and Moderna boosters respectively.

### Policy implications and interpretation

The effects of vaccination campaigns must be considered in the context in which they are implemented, due to changes in circulating variants, prevalence of previous infections, and changes in vaccine coverage. Our study was conducted during a time of high substantial pre-existing immunity. All participants had received at least two COVID-19 vaccine doses, with 85% having received three prior doses, and natural infection during successive omicron waves in 2022 was widespread. We previously estimated absolute vaccine effectiveness (compared with unvaccinated individuals) against COVID-19 hospitalisation of >80% at 6 months after primary vaccination with ChAdOx1-S among individuals aged 40–64 years in England, and this protection would have been enhanced among individuals who received a vaccine booster dose.(3) The resulting high baseline immunity in our study population may have reduced the effectiveness of the autumn 2022 booster campaign.

The autumn 2022 booster campaign in England was initiated to mitigate a probable winter COVID-19 wave. Vaccination campaigns are planned based on the best available knowledge at the time and cannot necessarily anticipate how widespread the wave will be. During our study period, the estimated prevalence of SARS-CoV-2 infection was between 2% and 3% among people 35-69 years, meaning that the risk of severe COVID-19 was low.(27) Our study estimated the impact of the autumn booster campaign in 50 year olds who were not clinically vulnerable, a group already at low risk of severe COVID-19. Based on the confidence intervals for the estimated effect, we cannot exclude a small reduction in the risk of severe outcomes. Booster vaccine effectiveness during autumn 2022 may have been higher in older adults, as was reported for booster doses administered in autumn 2021.(7)

There are other potential benefits of booster vaccination that we were not able to estimate. Illness that does not result in hospitalisation or death can still lead to loss of productivity and potentially long-term post-acute sequelae of COVID-19.(28,29) Further, vaccination of non-high risk individuals can indirectly benefit the rest of the community by limiting transmission. However, booster coverage was modest (40% in people aged 50.25 years). These findings may also not hold in future waves, with different SARS-CoV-2 transmissibility, circulating variants, and infection prevalence.

While COVID-19 booster vaccinations are extremely safe,(30) no pharmacological intervention is without risk. The relative risks and benefits of health interventions need to be considered, and the balance is likely to shift over time as the pandemic matures. For instance, in March 2023 the WHO Strategic Advisory Group of Experts on Immunization updated their guidance and does not recommend second boosters for non-high risk individuals, which includes healthy people aged <60 years.(31) Thus, there may be a case for changing eligibility thresholds in future campaigns, especially if vaccine costs and availability are a consideration, and there are expectations of low vaccine coverage. Continued monitoring of vaccination campaigns for emerging signals is warranted.

### Conclusion

In this study, we found little evidence that the autumn 2022 COVID-19 booster vaccination (coinciding with the influenza vaccination campaign) in England reduced severe COVID-19-related outcomes among non-clinically vulnerable 50-year-olds. We cannot rule out other impacts of the booster vaccination campaign on the 2022/23 winter COVID-19 wave, such as reducing virus transmission and reducing rates of mild or moderate COVID-19.

## Data Availability

Access to the underlying identifiable and potentially re-identifiable pseudonymised electronic health record data is tightly governed by various legislative and regulatory frameworks, and restricted by best practice. The data in OpenSAFELY is drawn from General Practice data across England where TPP is the Data Processor. TPP developers initiate an automated process to create pseudonymised records in the core OpenSAFELY database, which are copies of key structured data tables in the identifiable records. These are linked onto key external data resources that have also been pseudonymised via SHA-512 one-way hashing of NHS numbers using a shared salt. DataLab developers and PIs holding contracts with NHS England have access to the OpenSAFELY pseudonymised data tables as needed to develop the OpenSAFELY tools. These tools in turn enable researchers with OpenSAFELY Data Access Agreements to write and execute code for data management and data analysis without direct access to the underlying raw pseudonymised patient data, and to review the outputs of this code. All code for the full data management pipeline, from raw data to completed results for this analysis, and for the OpenSAFELY platform as a whole is available for review at github.com/OpenSAFELY.

## Acknowledgements

We are very grateful for all the support received from the TPP Technical Operations team throughout this work, and for generous assistance from the information governance and database teams at NHS England and the NHS England Transformation Directorate.

Members of the The OpenSAFELY Collaborative include: Sebastian CJ Bacon, Lucy Bridges, Benjamin FC Butler-Cole, Simon Davy, Iain Dillingham, David Evans, Ben Goldacre, Amelia Green, Liam Hart, George Hickman, Peter Inglesby, Steven Maude, Jessica Morley, Amir Mehrkar, Thomas O’Dwyer, Rebecca M Smith, Tom Ward, Jon Massey, Milan Wiedemann, Christopher Bates, Jonathan Cockburn, Sam Harper, Frank Hester, John Parry.

## Administrative

### Funding

The OpenSAFELY Platform is supported by grants from the Wellcome Trust (222097/Z/20/Z) and MRC (MR/V015757/1, MC_PC-20059, MR/W016729/1). In addition, development of OpenSAFELY has been funded by the Longitudinal Health and Wellbeing strand of the National Core Studies programme (MC_PC_20030: MC_PC_20059), the NIHR funded CONVALESCENCE programme (COV-LT-0009), NIHR (NIHR135559, COV-LT2-0073), and the Data and Connectivity National Core Study funded by UK Research and Innovation (MC_PC_20058), and Health Data Research UK (HDRUK2021.000, 2021.0157). EPKP is funded by the NIHR Health Protection Research Unit in Vaccines and Immunisation (NIHR200929), a partnership between the UK Health Security Agency and the London School of Hygiene & Tropical Medicine.

The views expressed are those of the authors and not necessarily those of the NIHR, NHS England, UK Health Security Agency (UKHSA) or the Department of Health and Social Care.

Funders had no role in the study design, collection, analysis, and interpretation of data; in the writing of the report; and in the decision to submit the article for publication.

### Information governance

NHS England is the data controller of the NHS England OpenSAFELY COVID-19 Service; TPP is the data processor; all study authors using OpenSAFELY have the approval of NHS England.(1) This implementation of OpenSAFELY is hosted within the TPP environment which is accredited to the ISO 27001 information security standard and is NHS IG Toolkit compliant.(2)

Patient data has been pseudonymised for analysis and linkage using industry standard cryptographic hashing techniques; all pseudonymised datasets transmitted for linkage onto OpenSAFELY are encrypted; access to the NHS England OpenSAFELY COVID-19 service is via a virtual private network (VPN) connection; the researchers hold contracts with NHS England and only access the platform to initiate database queries and statistical models; all database activity is logged; only aggregate statistical outputs leave the platform environment following best practice for anonymisation of results such as statistical disclosure control for low cell counts.(3)

The service adheres to the obligations of the UK General Data Protection Regulation (UK GDPR) and the Data Protection Act 2018. The service previously operated under notices initially issued in February 2020 by the Secretary of State under Regulation 3(4) of the Health Service (Control of Patient Information) Regulations 2002 (COPI Regulations), which required organisations to process confidential patient information for COVID-19 purposes; this set aside the requirement for patient consent.(4) As of 1 July 2023, the Secretary of State has requested that NHS England continue to operate the Service under the COVID-19 Directions 2020.(5) In some cases of data sharing, the common law duty of confidence is met using, for example, patient consent or support from the Health Research Authority Confidentiality Advisory Group.(6)

Taken together, these provide the legal bases to link patient datasets using the service. GP practices, which provide access to the primary care data, are required to share relevant health information to support the public health response to the pandemic, and have been informed of how the service operates.

(1) NHS Digital. The NHS England OpenSAFELY COVID-19 service - privacy notice [Internet]. 2023 [cited 2023 Jul 5]. Available from: https://digital.nhs.uk/coronavirus/coronavirus-covid-19-response-information-governance-hub/the-nhs-england-opensafely-covid-19-service-privacy-notice
(2) NHS Digital. NHS Digital. 2023 [cited 2023 Jul 5]. Data Security and Protection Toolkit. Available from: https://digital.nhs.uk/data-and-information/looking-after-information/data-security-and-information-governance/data-security-and-protection-toolkit
(3) NHS Digital [Internet]. [cited 2023 Mar 6]. ISB1523: Anonymisation Standard for Publishing Health and Social Care Data. Available from: https://digital.nhs.uk/data-and-information/information-standards/information-standards-and-data-collections-including-extractions/publications-and-notifications/standards-and-collections/isb1523-anonymisation-standard-for-publishing-health-and-social-care-data
(4) UK Department of Health and Social Care. GOV.UK. 2022 [cited 2023 Jul 5]. [Withdrawn] Coronavirus (COVID-19): notice under regulation 3(4) of the Health Service (Control of Patient Information) Regulations 2002 – general. Available from: https://www.gov.uk/government/publications/coronavirus-covid-19-notification-of-data-controllers-to-share-information/coronavirus-covid-19-notice-under-regulation-34-of-the-health-service-control-of-patient-information-regulations-2002-general--2
(5) NHS Digital. NHS Digital. 2022 [cited 2023 Jul 5]. Secretary of State for Health and Social Care: COVID-19 Public Health Directions 2020. Available from: https://digital.nhs.uk/about-nhs-digital/corporate-information-and-documents/directions-and-data-provision-notices/secretary-of-state-directions/covid-19-public-health-directions-2020
(6) NHS Health Research Authority. Health Research Authority. [cited 2023 Jul 5]. Confidentiality Advisory Group. Available from: https://www.hra.nhs.uk/about-us/committees-and-services/confidentiality-advisory-group/

## Supplementary files

**Supplementary Table 1.** List of exclusion criteria and their definition. Unless otherwise stated, all criteria were identified using primary care data. Codelsits with all codes used to define criteria area available in the Github repository: https://github.com/opensafely/vax-fourth-dose-RD

| Exclusion criteria |  | Definition |
| --- | --- | --- |
| JCVI high risk groups prioritised for vaccination(1) | Chronic respiratory disease (including poorly controlled asthma) | - Chronic respiratory disease diagnosis anytime prior to index date;<br>- Asthma admission recorded in primary care anytime prior to index date; OR<br>- Asthma diagnosis anytime prior to index date with a prescription for systemic steroids in each of the 3 months prior to the index date. |
|  | Chronic heart disease and vascular disease | Diagnosis anytime prior to index date |
|  | Chronic kidney disease | Diagnosis anytime prior to index date |
|  | Chronic liver disease | Diagnosis anytime prior to index date |
|  | Chronic neurological disease | Diagnosis anytime prior to index date |
|  | Learning disability | Diagnosis anytime prior to index date |
|  | Diabetes mellitus and other endocrine disorders | Diagnosis anytime prior to index date, either in the absence, or occurring after, a resolved diabetes code |
|  | Immunosuppressed (including HIV/AIDS) | - Diagnosis of condition causing immunosuppression (including HIV infection/AIDS or solid organ transplant) anytime prior to index date; OR<br>- Cancer diagnosis anytime in 3 years prior to index date; OR<br>- Prescription for a chemotherapeutic, immunosuppressant, or immunomodulating medicine in 6 months prior to the index date |
|  | Morbid obesity | - Diagnosis of severe obesity (BMI $\geq 40$) following date of BMI being recorded; OR<br>- Most recent recorded BMI value $\geq 40$ |
|  | Asplenia | Diagnosis anytime prior to index date |
|  | Severe mental illness | Diagnosis anytime prior to index date, either in the absence, or occurring after, a severe mental illness in remission code |
| Receipt of third or fourth booster dose prior to first availability to non-immunosuppressed population |  | Evidence of a third COVID-19 vaccination prior to 16 September 2021(2), or a fourth COVID-19 vaccination prior to 5 September 2022(3) |
| Health or social care worker |  | Stated that they were a health or social care worker when receiving at least one of their COVID-19 vaccinations |
| End of life |  | - Code indicating end of life recorded in primary care; OR<br>- Prescription for midazolam injection indicated for treatment of pain at end of life any time prior to index date. |
| Residents in a care or nursing home |  | - Care home residence code anytime prior to index date; OR<br>- Current address maps to list of care homes(4) |
| Housebound people |  | - Code indicating person is housebound, in absence of: a code indicating that the person is no longer housebound; a code indicating the person is in a care home |

(1) UK Health Security Agency. GOV.UK. 2023 [cited 2023 Mar 10]. COVID-19: the green book, chapter 14a. Available from: https://www.gov.uk/government/publications/covid-19-the-green-book-chapter-14a
(2) NHS England. NHS begins COVID-19 booster vaccination campaign [Internet]. [cited 2023 Mar 17]. Available from: https://www.england.nhs.uk/2021/09/nhs-begins-covid-19-booster-vaccination-campaign/
(3) NHS England. NHS invites people 50 and over for autumn boosters and flu jab [Internet]. 2022 [cited 2023 Mar 10]. Available from: https://www.england.nhs.uk/2022/10/nhs-invites-people-50-and-over-for-autumn-boosters-and-flu-jab/
(4) Schultze A, Bates C, Cockburn J, MacKenna B, Nightingale E, Curtis HJ, et al. Identifying Care Home Residents in Electronic Health Records - An OpenSAFELY Short Data Report. Wellcome Open Res. 2021;(6):90.

**Supplementary Table 2.** List of codes used to define outcomes.

| **Outcome** | **Codelist** |
| --- | --- |
| **COVID-19 hospital admission** | Unplanned hospital admission with any of the following ICD-10 codes in any position (primary or secondary): U071, U072, U099, U109 |
| **COVID-19 accident and emergency attendance** | A&E attendances with any of the following SNOMED codes: 1240751000000100, 1325161000000102, 1325171000000109, 132581000000106 |
| **COVID-19 death** | Deaths with any of the following ICD-10 codes on the death certificate in any position (underlying or contributing): U071, U072, U099, U109 |
| **Respiratory admission** | Unplanned hospital admission with any of the following ICD-10 codes in the primary position (J00-J99) |
| **Respiratory death** | Deaths with any of the following ICD-10 codes on the death certificate (underlying only): (J00-J99) |

**Supplementary Figure 1.**
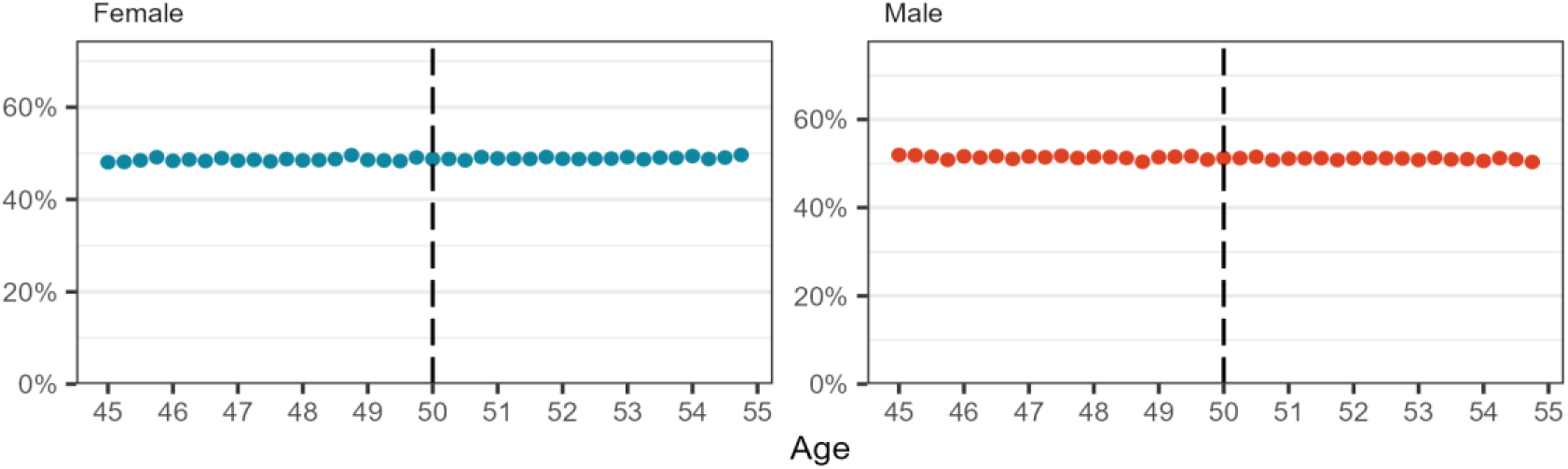
Sex relative frequency distribution by age in 3-month intervals based on age at 3 September 2022.

**Supplementary Figure 2.**
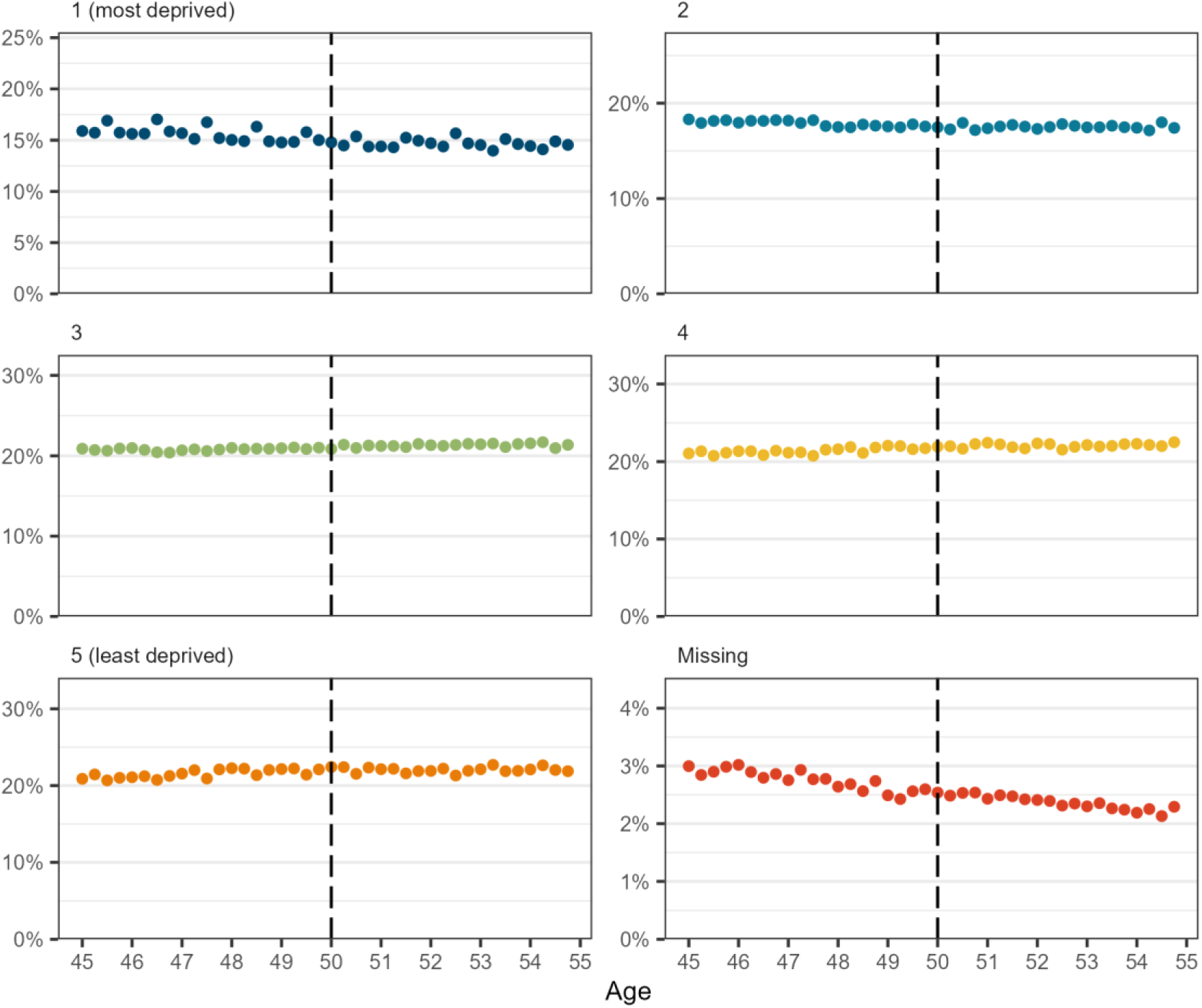
IMD relative frequency distribution by age in 3-month intervals based on age at 3 September 2022

**Supplementary Figure 3.**
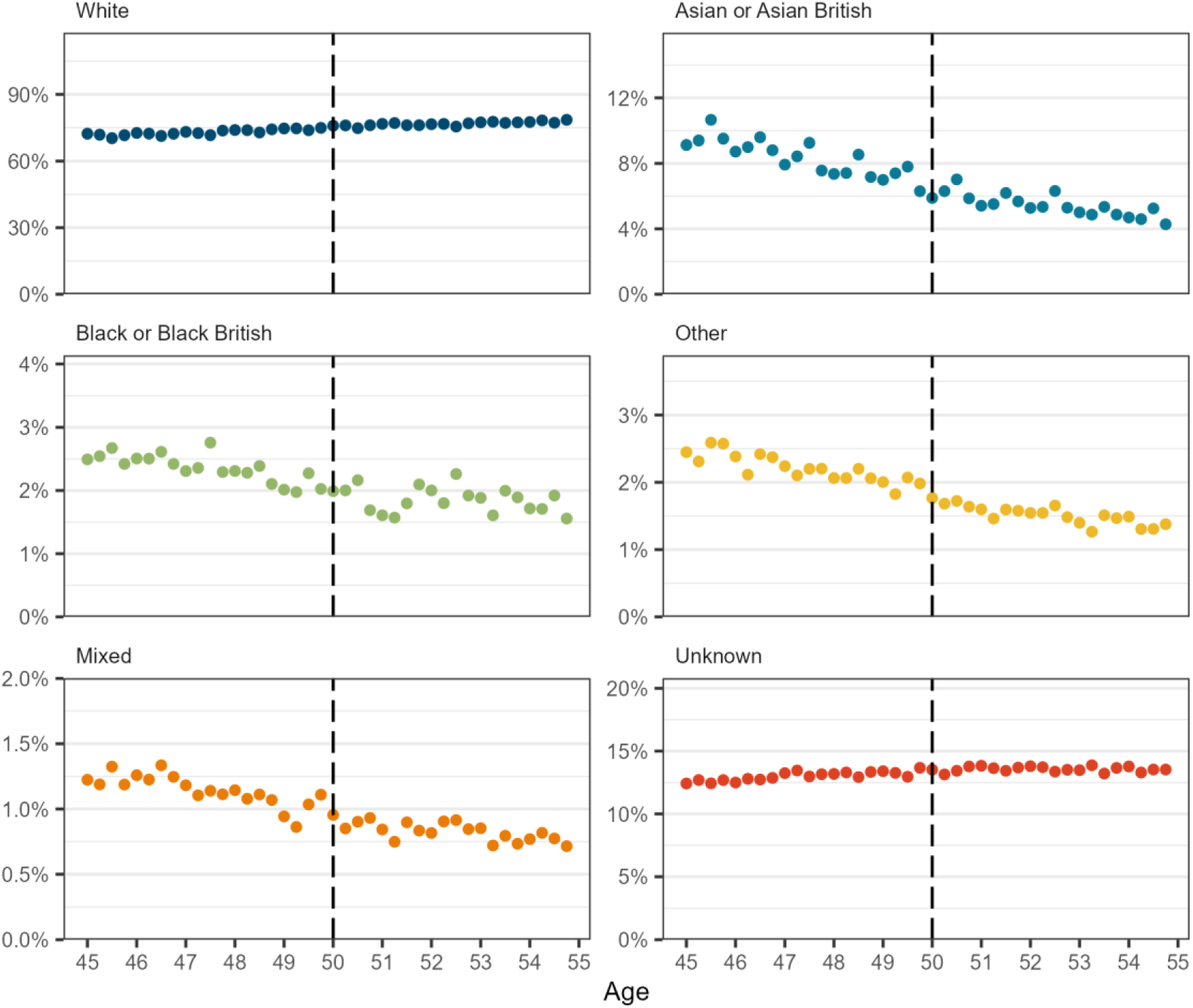
Ethnicity relative frequency distribution by age in 3-month intervals based on age at 3 September 2022

**Supplementary Figure 4.**
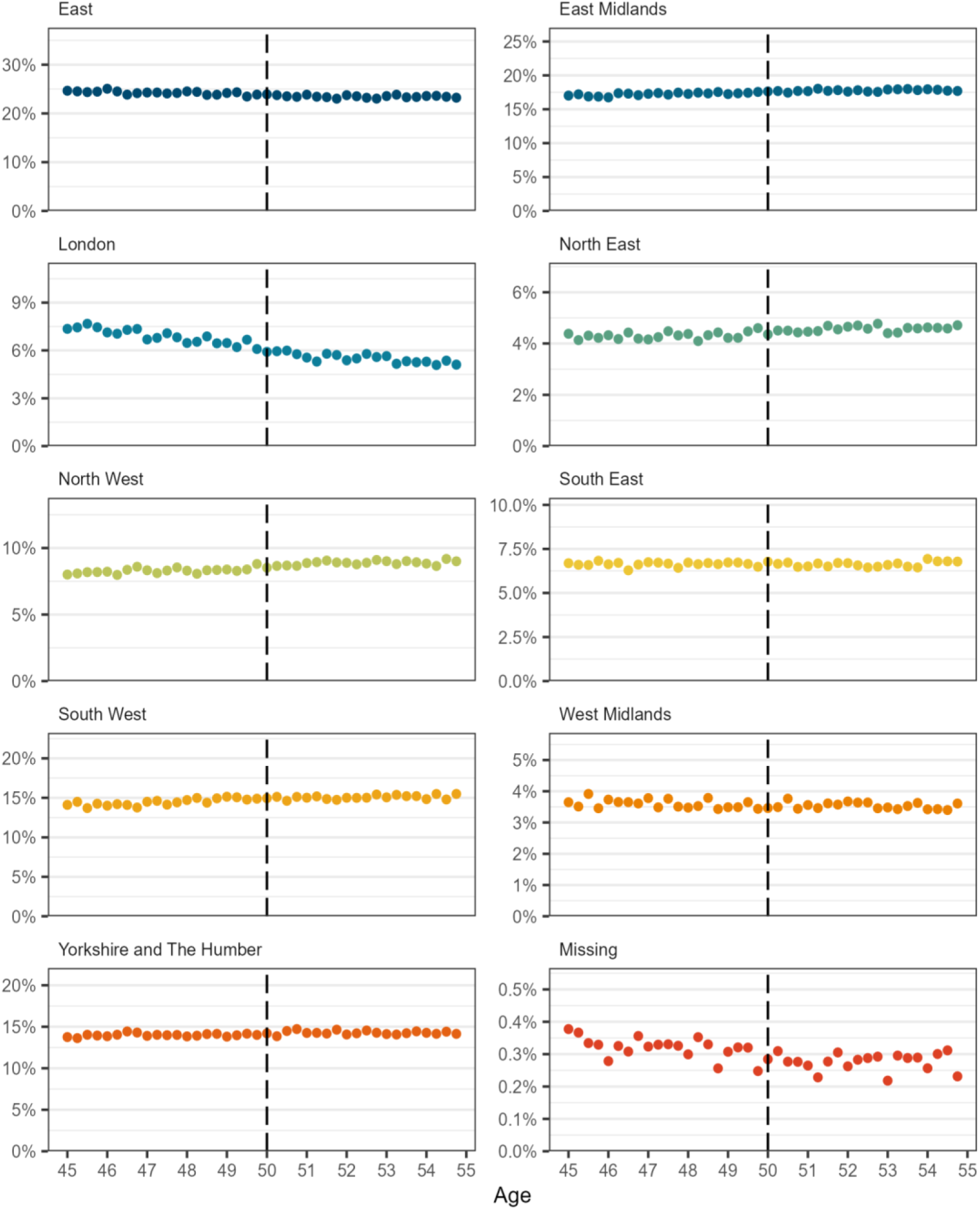
Region relative frequency distribution by age in 3-month intervals based on age at 3 September 2022

**Supplementary Figure 5.**
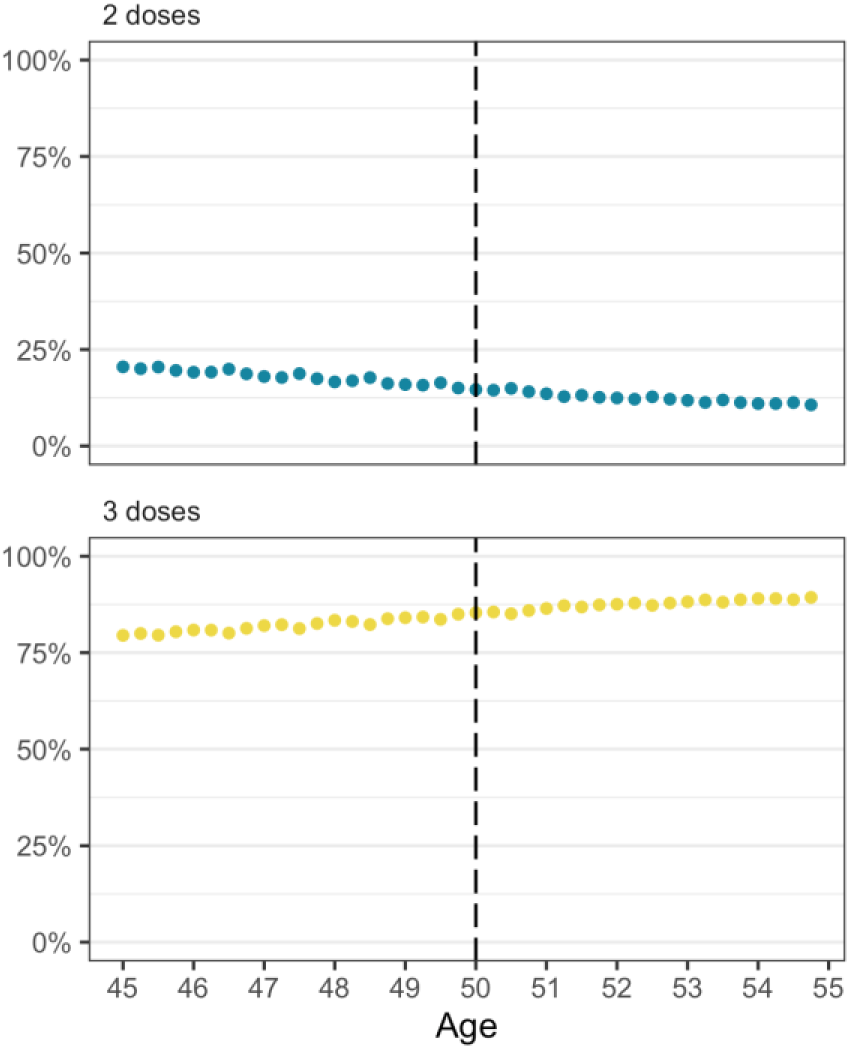
Relative frequency distribution of previous number of COVID-19 vaccine doses by age in 3-month intervals based on age at 3 September 2022

**Supplementary Figure 6.**
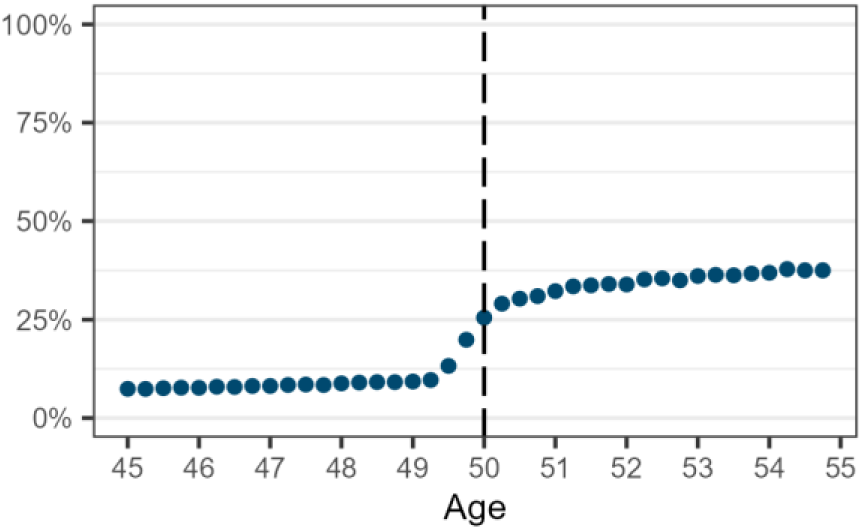
Coverage of 2022/23 influenza vaccination by 26 November 2023 by age in 3-month intervals

**Supplementary Table 3.**
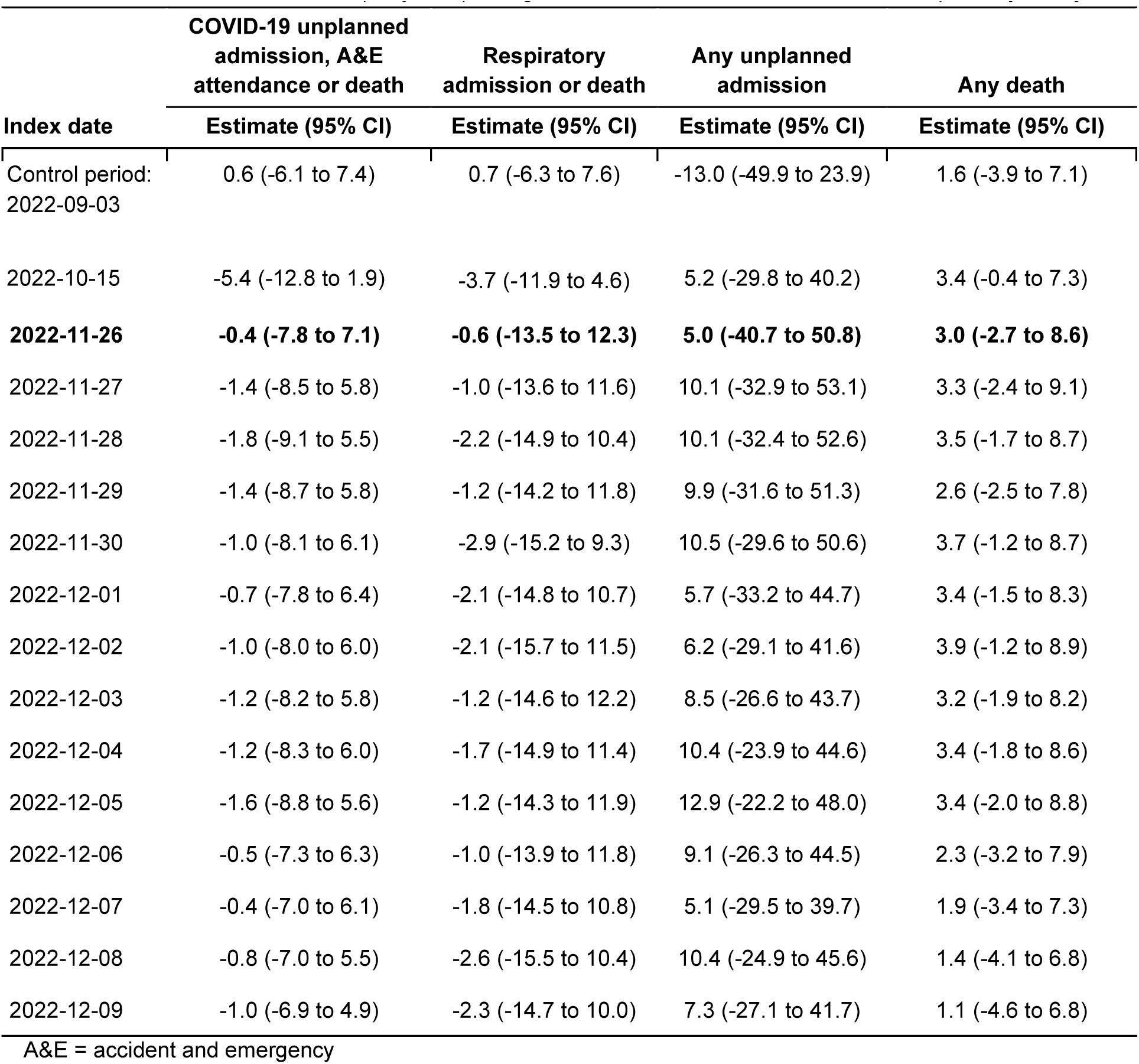
Estimates from sharp regression discontinuity analysis estimating change in 6-week outcomes at threshold (50 years) using different index dates. Bold indicates primary analysis.

| Index date | COVID-19 unplanned admission, A&E attendance or death | Respiratory admission or death | Any unplanned admission | Any death |
| --- | --- | --- | --- | --- |
|  | Estimate (95% CI) | Estimate (95% CI) | Estimate (95% CI) | Estimate (95% CI) |
| Control period:<br>2022-09-03 | 0.6 (-6.1 to 7.4) | 0.7 (-6.3 to 7.6) | -13.0 (-49.9 to 23.9) | 1.6 (-3.9 to 7.1) |
| 2022-10-15 | -5.4 (-12.8 to 1.9) | -3.7 (-11.9 to 4.6) | 5.2 (-29.8 to 40.2) | 3.4 (-0.4 to 7.3) |
| **2022-11-26** | **-0.4 (-7.8 to 7.1)** | **-0.6 (-13.5 to 12.3)** | **5.0 (-40.7 to 50.8)** | **3.0 (-2.7 to 8.6)** |
| 2022-11-27 | -1.4 (-8.5 to 5.8) | -1.0 (-13.6 to 11.6) | 10.1 (-32.9 to 53.1) | 3.3 (-2.4 to 9.1) |
| 2022-11-28 | -1.8 (-9.1 to 5.5) | -2.2 (-14.9 to 10.4) | 10.1 (-32.4 to 52.6) | 3.5 (-1.7 to 8.7) |
| 2022-11-29 | -1.4 (-8.7 to 5.8) | -1.2 (-14.2 to 11.8) | 9.9 (-31.6 to 51.3) | 2.6 (-2.5 to 7.8) |
| 2022-11-30 | -1.0 (-8.1 to 6.1) | -2.9 (-15.2 to 9.3) | 10.5 (-29.6 to 50.6) | 3.7 (-1.2 to 8.7) |
| 2022-12-01 | -0.7 (-7.8 to 6.4) | -2.1 (-14.8 to 10.7) | 5.7 (-33.2 to 44.7) | 3.4 (-1.5 to 8.3) |
| 2022-12-02 | -1.0 (-8.0 to 6.0) | -2.1 (-15.7 to 11.5) | 6.2 (-29.1 to 41.6) | 3.9 (-1.2 to 8.9) |
| 2022-12-03 | -1.2 (-8.2 to 5.8) | -1.2 (-14.6 to 12.2) | 8.5 (-26.6 to 43.7) | 3.2 (-1.9 to 8.2) |
| 2022-12-04 | -1.2 (-8.3 to 6.0) | -1.7 (-14.9 to 11.4) | 10.4 (-23.9 to 44.6) | 3.4 (-1.8 to 8.6) |
| 2022-12-05 | -1.6 (-8.8 to 5.6) | -1.2 (-14.3 to 11.9) | 12.9 (-22.2 to 48.0) | 3.4 (-2.0 to 8.8) |
| 2022-12-06 | -0.5 (-7.3 to 6.3) | -1.0 (-13.9 to 11.8) | 9.1 (-26.3 to 44.5) | 2.3 (-3.2 to 7.9) |
| 2022-12-07 | -0.4 (-7.0 to 6.1) | -1.8 (-14.5 to 10.8) | 5.1 (-29.5 to 39.7) | 1.9 (-3.4 to 7.3) |
| 2022-12-08 | -0.8 (-7.0 to 5.5) | -2.6 (-15.5 to 10.4) | 10.4 (-24.9 to 45.6) | 1.4 (-4.1 to 6.8) |
| 2022-12-09 | -1.0 (-6.9 to 4.9) | -2.3 (-14.7 to 10.0) | 7.3 (-27.1 to 41.7) | 1.1 (-4.6 to 6.8) |

A&E = accident and emergency

**Supplementary Table 4.** Estimates from sharp regression discontinuity analysis excluding people born in the index month, estimating change in 6-week outcomes at threshold (50 years) using different index dates. Bold indicates primary analysis.

| Index date | COVID-19 unplanned admission, A&E attendance or death | Respiratory admission or death | Any unplanned admission | Any death |
| --- | --- | --- | --- | --- |
|  | Estimate (95% CI) | Estimate (95% CI) | Estimate (95% CI) | Estimate (95% CI) |
| Control period: 2022-09-03 | 1.3 (-5.9 to 8.4) | -2.1 (-8.8 to 4.5) | -16.0 (-55.1 to 23.1) | 2.5 (-3.2 to 8.3) |
| 2022-10-15 | -5.8 (-13.6 to 2.0) | -4.8 (-13.4 to 3.9) | 1.3 (-35.7 to 38.3) | 2.7 (-1.4 to 6.7) |
| **2022-11-26** | **-1.8 (-9.5 to 6.0)** | **-2.8 (-16.2 to 10.7)** | **-3.6 (-51.1 to 43.9)** | **4.6 (-1.1 to 10.3)** |
| 2022-11-27 | -2.8 (-10.2 to 4.6) | -3.2 (-16.3 to 9.9) | 2.4 (-42.4 to 47.1) | 4.9 (-0.9 to 10.8) |
| 2022-11-28 | -3.2 (-10.8 to 4.3) | -4.2 (-17.4 to 9.0) | 1.3 (-42.6 to 45.2) | 5.2 (0.0 to 10.4) |
| 2022-11-29 | -3.2 (-10.7 to 4.2) | -2.5 (-16.2 to 11.2) | 0.9 (-41.8 to 43.6) | 4.2 (-0.9 to 9.4) |
| 2022-11-30 | -2.8 (-10.0 to 4.5) | -4.3 (-17.2 to 8.7) | 2.6 (-38.9 to 44.2) | 5.1 (0.0 to 10.1) |
| 2022-12-01 | -2.5 (-9.7 to 4.8) | -3.7 (-17.1 to 9.7) | -3.4 (-43.3 to 36.6) | 4.7 (-0.3 to 9.6) |
| 2022-12-02 | -2.8 (-10.0 to 4.3) | -3.9 (-18.2 to 10.4) | -0.5 (-37.2 to 36.2) | 5.2 (0.0 to 10.3) |
| 2022-12-03 | -3.0 (-10.2 to 4.1) | -3.1 (-17.2 to 11.1) | 2.0 (-34.5 to 38.6) | 4.8 (-0.1 to 9.8) |
| 2022-12-04 | -3.1 (-10.4 to 4.1) | -3.7 (-17.4 to 10.1) | 3.5 (-32.0 to 38.9) | 5.1 (-0.1 to 10.2) |
| 2022-12-05 | -3.6 (-10.9 to 3.7) | -3.6 (-17.2 to 10.0) | 5.2 (-30.9 to 41.4) | 5.1 (-0.3 to 10.4) |
| 2022-12-06 | -2.4 (-9.3 to 4.4) | -2.4 (-15.9 to 11.2) | 2.3 (-34.4 to 39.1) | 3.9 (-1.6 to 9.5) |
| 2022-12-07 | -2.4 (-8.9 to 4.1) | -2.2 (-15.7 to 11.2) | -0.9 (-37.0 to 35.1) | 3.5 (-1.8 to 8.8) |
| 2022-12-08 | -2.3 (-8.7 to 4.1) | -2.1 (-15.8 to 11.7) | 2.6 (-33.7 to 38.9) | 2.9 (-2.5 to 8.4) |
| 2022-12-09 | -2.2 (-8.3 to 3.9) | -1.5 (-14.5 to 11.6) | -1.1 (-36.2 to 34.0) | 2.6 (-3.2 to 8.3) |

A&E = accident and emergency

**Supplementary Table 5.** Estimates from sharp regression discontinuity analysis estimating change in 6-week outcomes at threshold (50 years) using different bandwidths for primary index date (26 November 2022).

|  | **COVID-19 unplanned admission, A&E attendance or death** | **Respiratory admission or death** | **Any unplanned admission** | **Any death** |
| --- | --- | --- | --- | --- |
| **Bandwidth** | **Estimate (95% CI)** | **Estimate (95% CI)** | **Estimate (95% CI)** | **Estimate (95% CI)** |
| 4 years | 1.0 (-7.0 to 9.1) | 2.4 (-12.3 to 17.1) | 30.0 (-22.3 to 82.2) | 1.3 (-4.7 to 7.3) |
| 3 years | -0.8 (-10.4 to 8.7) | 2.9 (-15.5 to 21.3) | 16.5 (-42.7 to 75.7) | 1.9 (-6.3 to 10.0) |
| 2 years | 1.9 (-9.5 to 13.3) | 2.2 (-19.7 to 24.0) | 16.3 (-62.9 to 95.5) | -5.9 (-15.7 to 3.8) |
| 1 year | 2.0 (-17.9 to 21.9) | 12.2 (-32.1 to 56.5) | 13.0 (-186.9 to 212.8) | -9.6 (-27.8 to 8.5) |

A&E = accident and emergency

**Supplementary Table 6.** Instrumental variable analysis (fuzzy regression discontinuity) estimating local average treatment effect (LATE) for 6-week outcomes with different index dates. Bold indicates primary analysis.

| Index date | COVID-19 unplanned admission, A&E attendance or death | Respiratory admission or death | Any unplanned admission | Any death |
| --- | --- | --- | --- | --- |
|  | Estimate (95% CI) | Estimate (95% CI) | Estimate (95% CI) | Estimate (95% CI) |
| **2022-11-26** | **2.1 (-11.3 to 15.4)** | **-12.0 (-58.2 to 34.3)** | **15.4 (-166.7 to 197.6)** | **14.7 (-6.4 to 35.8)** |
| 2022-11-27 | -1.4 (-14.4 to 11.7) | -12.3 (-54.4 to 29.7) | 32.8 (-128.0 to 193.5) | 16.0 (-3.6 to 35.6) |
| 2022-11-28 | -2.8 (-16.3 to 10.8) | -17.1 (-58.6 to 24.4) | 30.9 (-130.1 to 191.8) | 16.7 (-1.6 to 35.0) |
| 2022-11-29 | -4.4 (-27.6 to 18.8) | -4.7 (-48.3 to 38.9) | 29.6 (-120.2 to 179.3) | 9.5 (-10.8 to 29.7) |
| 2022-11-30 | -2.7 (-24.6 to 19.1) | -10.4 (-51.3 to 30.4) | 31.8 (-112.4 to 176.0) | 13.2 (-6.3 to 32.7) |
| 2022-12-01 | -2.4 (-25.2 to 20.3) | -6.9 (-54.5 to 40.6) | 19.7 (-125.6 to 165.0) | 11.5 (-8.5 to 31.5) |
| 2022-12-02 | -3.5 (-26.7 to 19.7) | -7.1 (-57.9 to 43.6) | 21.1 (-118.4 to 160.6) | 13.0 (-7.8 to 33.8) |
| 2022-12-03 | -3.9 (-27.1 to 19.2) | -4.0 (-54.7 to 46.7) | 28.6 (-102.9 to 160.1) | 10.6 (-10.6 to 31.8) |
| 2022-12-04 | -3.8 (-29.2 to 21.5) | -5.7 (-55.6 to 44.2) | 34.4 (-99.2 to 168.0) | 11.3 (-9.3 to 31.8) |
| 2022-12-05 | -5.3 (-30.7 to 20.1) | -3.9 (-54.1 to 46.4) | 42.6 (-92.1 to 177.3) | 11.1 (-9.7 to 31.9) |
| 2022-12-06 | -1.7 (-25.1 to 21.7) | -3.4 (-51.3 to 44.5) | 29.9 (-106.1 to 165.9) | 7.6 (-12.9 to 28.1) |
| 2022-12-07 | -1.5 (-24.5 to 21.6) | -6.0 (-51.3 to 39.4) | 16.8 (-107.1 to 140.8) | 6.3 (-12.9 to 25.5) |
| 2022-12-08 | -2.5 (-24.2 to 19.2) | -8.3 (-55.3 to 38.7) | 33.7 (-97.9 to 165.2) | 4.4 (-15.3 to 24.1) |
| 2022-12-09 | -3.3 (-23.4 to 16.8) | -7.5 (-55.0 to 40.1) | 23.7 (-108.7 to 156.2) | 3.5 (-16.4 to 23.4) |

A&E = accident and emergency

**Supplementary Table 7.** Instrumental variable analysis (fuzzy regression discontinuity) estimating local average treatment effect (LATE) for 6-week outcomes with different index dates including receipt of influenza vaccination in the model. Bold indicates primary analysis.

| Index date | COVID-19 unplanned admission, A&E attendance or death | Respiratory admission or death | Any unplanned admission | Any death |
| --- | --- | --- | --- | --- |
|  | Estimate (95% CI) | Estimate (95% CI) | Estimate (95% CI) | Estimate (95% CI) |
| **2022-11-26** | **5.4 (-24.9 to 35.8)** | **-24.7 (-153.9 to 104.6)** | **8.3 (-286.4 to 303.1)** | **27.1 (-10.5 to 64.8)** |
| 2022-11-27 | 0.8 (-30.1 to 31.7) | -24.1 (-145.9 to 97.8) | 36.3 (-251.8 to 324.4) | 28.6 (-7.3 to 64.5) |
| 2022-11-28 | -1.7 (-39.0 to 35.7) | -29.9 (-148.0 to 88.1) | 29.5 (-267.6 to 326.6) | 29.7 (-7.4 to 66.9) |
| 2022-11-29 | -4.0 (-47.1 to 39.1) | -14.9 (-131.7 to 101.9) | 24.3 (-268.2 to 316.8) | 19.7 (-28.6 to 68.1) |
| 2022-11-30 | -2.0 (-42.7 to 38.7) | -24.1 (-154.9 to 106.7) | 26.8 (-281 to 334.5) | 25.1 (-22.3 to 72.4) |
| 2022-12-01 | -2.0 (-44.6 to 40.6) | -19.2 (-157.9 to 119.5) | 5.7 (-298.2 to 309.5) | 22.7 (-18.2 to 63.6) |
| 2022-12-02 | -3.5 (-45.3 to 38.2) | -18.2 (-159.4 to 123.0) | 8.5 (-283.7 to 300.6) | 24.9 (-17.3 to 67.1) |
| 2022-12-03 | -4.1 (-45.2 to 36.9) | -15.0 (-156.6 to 126.5) | 13.0 (-263.3 to 289.2) | 21.3 (-17.3 to 59.8) |
| 2022-12-04 | -4.2 (-47.1 to 38.7) | -17.5 (-164.1 to 129.1) | 22.6 (-272.7 to 317.9) | 22.1 (-13.8 to 58.0) |
| 2022-12-05 | -6.8 (-48.9 to 35.3) | -15.0 (-163.3 to 133.3) | 31.0 (-265.8 to 327.8) | 22.1 (-14.4 to 58.5) |
| 2022-12-06 | -1.9 (-43.5 to 39.7) | -13.5 (-162.2 to 135.1) | 14.0 (-278.9 to 306.9) | 16.9 (-18.0 to 51.7) |
| 2022-12-07 | -2.1 (-42.1 to 37.9) | -17.0 (-163.8 to 129.7) | -7.9 (-293.8 to 277.9) | 14.9 (-17.9 to 47.7) |
| 2022-12-08 | -3.5 (-41.7 to 34.7) | -19.1 (-163.2 to 125.0) | 16.4 (-291.5 to 324.3) | 12.1 (-20.3 to 44.5) |
| 2022-12-09 | -4.6 (-40.0 to 30.8) | -17.6 (-157.4 to 122.1) | 2.7 (-308.6 to 314.1) | 10.9 (-21.4 to 43.1) |

A&E = accident and emergency

**Supplementary Table 8.** Estimates from fuzzy regression discontinuity analysis estimating change in 6-week outcomes at threshold (50 years) using different bandwidths for primary index date (26 November 2022).

|  | **COVID-19 unplanned admission, A&E attendance or death** | **Respiratory admission or death** | **Any unplanned admission** | **Any death** |
| --- | --- | --- | --- | --- |
| **Bandwidth** | **Estimate (95% CI)** | **Estimate (95% CI)** | **Estimate (95% CI)** | **Estimate (95% CI)** |
| 4 years | 5.5 (-9.9 to 21.0) | -0.8 (-52.8 to 51.2) | 92.0 (-128.0 to 311.9) | 12.6 (-12.4 to 37.7) |
| 3 years | -5.7 (-26.6 to 15.3) | -3.0 (-69.3 to 63.3) | 51.2 (-218 to 320.4) | 10.4 (-18.9 to 39.7) |
| 2 years | 0.6 (-22.0 to 23.2) | -1.0 (-75.5 to 73.4) | 38.6 (-290.2 to 367.5) | -12.0 (-46.7 to 22.8) |
| 1 year | 5.5 (-9.9 to 21.0) | -0.8 (-52.8 to 51.2) | 92.0 (-128.0 to 311.9) | 12.6 (-12.4 to 37.7) |

A&E = accident and emergency

## References

1. Polack FP, Thomas SJ, Kitchin N, Absalon J, Gurtman A, Lockhart S, et al. Safety and Efficacy of the BNT162b2 mRNA Covid-19 Vaccine. N Engl J Med. 2020 Dec 31;383(27):2603–15.

2. Voysey M, Clemens SAC, Madhi SA, Weckx LY, Folegatti PM, Aley PK, et al. Safety and efficacy of the ChAdOx1 nCoV-19 vaccine (AZD1222) against SARS-CoV-2: an interim analysis of four randomised controlled trials in Brazil, South Africa, and the UK. The Lancet. 2021 Jan 9;397(10269):99–111.

3. Horne EMF, Hulme WJ, Keogh RH, Palmer TM, Williamson EJ, Parker EPK, et al. Waning effectiveness of BNT162b2 and ChAdOx1 covid-19 vaccines over six months since second dose: OpenSAFELY cohort study using linked electronic health records. BMJ. 2022 Jul 20;378:e071249.

4. Kirsebom FCM, Andrews N, Stowe J, Toffa S, Sachdeva R, Gallagher E, et al. COVID-19 vaccine effectiveness against the omicron (BA.2) variant in England. Lancet Infect Dis. 2022 Jul 1;22(7):931–3.

5. Andrews N, Stowe J, Kirsebom F, Toffa S, Rickeard T, Gallagher E, et al. Covid-19 Vaccine Effectiveness against the Omicron (B.1.1.529) Variant. N Engl J Med. 2022 Apr 21;386(16):1532–46.

6. Stowe J, Andrews N, Kirsebom F, Ramsay M, Bernal JL. Effectiveness of COVID-19 vaccines against Omicron and Delta hospitalisation, a test negative case-control study. Nat Commun. 2022 Sep 30;13(1):5736.

7. Hulme WJ, Williamson EJ, Horne E, Green A, Nab L, Keogh R, et al. Effectiveness of BNT162b2 booster doses in England: an observational study in OpenSAFELY-TPP [Internet]. medRxiv; 2022 [cited 2023 Jun 20]. p. 2022.06.06.22276026. Available from: https://www.medrxiv.org/content/10.1101/2022.06.06.22276026v1

8. Arbel R, Hammerman A, Sergienko R, Friger M, Peretz A, Netzer D, et al. BNT162b2 Vaccine Booster and Mortality Due to Covid-19. N Engl J Med. 2021 Dec 23;385(26):2413–20.

9. Andrews N, Stowe J, Kirsebom F, Toffa S, Sachdeva R, Gower C, et al. Effectiveness of COVID-19 booster vaccines against COVID-19-related symptoms, hospitalization and death in England. Nat Med. 2022 Apr;28(4):831–7.

10. NHS England. NHS COVID-19 vaccine bookings to open to millions of people as autumn booster campaign kicks off [Internet]. 2022 [cited 2023 Mar 10]. Available from: https://www.england.nhs.uk/2022/09/vaccine-bookings-to-open-to-millions-of-people-as-autumn-booster-campaign-kicks-off/

11. NHS England. Over 65s can now book autumn COVID booster [Internet]. 2022 [cited 2023 Mar 10]. Available from: https://www.england.nhs.uk/2022/09/over-65s-can-now-book-autumn-covid-booster/

12. NHS England. NHS invites people 50 and over for autumn boosters and flu jab [Internet]. 2022 [cited 2023 Mar 10]. Available from: https://www.england.nhs.uk/2022/10/nhs-invites-people-50-and-over-for-autumn-boosters-and-flu-jab/

13. UK Health Security Agency. GOV.UK. 2023 [cited 2023 Mar 10]. COVID-19: the green book, chapter 14a. Available from: https://www.gov.uk/government/publications/covid-19-the-green-book-chapter-14a

14. Hulme WJ, Horne EMF, Parker EPK, Keogh RH, Williamson EJ, Walker V, et al. Comparative effectiveness of BNT162b2 versus mRNA-1273 covid-19 vaccine boosting in England: matched cohort study in OpenSAFELY-TPP. BMJ. 2023 Mar 15;380:e072808.

15. Oldenburg CE, Moscoe E, Bärnighausen T. Regression Discontinuity for Causal Effect Estimation in Epidemiology. Curr Epidemiol Rep. 2016;3:233.

16. Bermingham C, Morgan J, Ayoubkhani D, Glickman M, Islam N, Sheikh A, et al. Estimating the Effectiveness of First Dose of COVID-19 Vaccine Against Mortality in England: A Quasi-Experimental Study. Am J Epidemiol. 2023 Feb 1;192(2):267–75.

17. Anderson ML, Dobkin C, Gorry D. The Effect of Influenza Vaccination for the Elderly on Hospitalization and Mortality. Ann Intern Med. 2020 Apr 7;172(7):445–52.

18. Moscoe E, Bor J, Bärnighausen T. Regression discontinuity designs are underutilized in medicine, epidemiology, and public health: a review of current and best practice. J Clin Epidemiol. 2015 Feb 1;68(2):132–43.

19. Smith LM, Lévesque LE, Kaufman JS, Strumpf EC. Strategies for evaluating the assumptions of the regression discontinuity design: a case study using a human papillomavirus vaccination programme. Int J Epidemiol. 2017 Jun 1;46(3):939–49.

20. Hernán MA, Robins JM. Causal Inference: What If [Internet]. Boca Raton: Chapman & Hall/CRC; 2020. Available from: https://www.hsph.harvard.edu/miguel-hernan/causal-inference-book/

21. Andrews C, Schultze A, Curtis H, Hulme W, Tazare J, Evans S, et al. OpenSAFELY: Representativeness of electronic health record platform OpenSAFELY-TPP data compared to the population of England. Wellcome Open Res. 2022;7:191.

22. Hulme WJ, Williamson E, Horne EMF, Green A, McDonald HI, Walker AJ, et al. Challenges in Estimating the Effectiveness of COVID-19 Vaccination Using Observational Data. Ann Intern Med. 2023 May 16;176(5):685–93.

23. Bar-On YM, Goldberg Y, Mandel M, Bodenheimer O, Amir O, Freedman L, et al. Protection by a Fourth Dose of BNT162b2 against Omicron in Israel. N Engl J Med. 2022 May 5;386(18):1712–20.

24. Mateo-Urdiales A, Sacco C, Fotakis EA, Manso MD, Bella A, Riccardo F, et al. Relative effectiveness of monovalent and bivalent mRNA boosters in preventing severe COVID-19 due to omicron BA.5 infection up to 4 months post-administration in people aged 60 years or older in Italy: a retrospective matched cohort study. Lancet Infect Dis. 2023 Jul 18;S1473-3099(23)00374–2.

25. Lin DY, Xu Y, Gu Y, Zeng D, Wheeler B, Young H, et al. Effectiveness of Bivalent Boosters against Severe Omicron Infection. N Engl J Med. 2023 Feb 23;388(8):764–6.

26. Kirsebom FCM, Andrews N, Stowe J, Ramsay M, Bernal JL. Duration of protection of ancestral-strain monovalent vaccines and effectiveness of bivalent BA.1 boosters against COVID-19 hospitalisation in England: a test-negative case-control study. Lancet Infect Dis. 2023 Jul 2023 Jul 12;S1473-3099(23)00365–1.

27. Office for National Statistics. Coronavirus (COVID-19) latest insights [Internet]. 2023 [cited 2023 Mar 31]. Available from: https://www.ons.gov.uk/peoplepopulationandcommunity/healthandsocialcare/conditionsanddiseases/articles/coronaviruscovid19latestinsights/infections

28. Thompson EJ, Williams DM, Walker AJ, Mitchell RE, Niedzwiedz CL, Yang TC, et al. Long COVID burden and risk factors in 10 UK longitudinal studies and electronic health records. Nat Commun. 2022 Jun 28;13(1):3528.

29. The OpenSAFELY Collaborative, Schaffer AL, Park RY, Tazare J, Bhaskaran K, MacKenna B, et al. Fit notes associated with COVID-19 in 24 million patients’ primary care records: A cohort study in OpenSAFELY-TPP. medRxiv 2023.07.28.23293269

30. Hause AM. Safety Monitoring of Bivalent COVID-19 mRNA Vaccine Booster Doses Among Persons Aged ≥12 Years — United States, August 31–October 23, 2022. MMWR Morb Mortal Wkly Rep. 2022 Nov 4;71(44):1401–1406.

31. World Health Organisation Strategic Advisory Group of Experts on Immunisation. SAGE updates COVID-19 vaccination guidance [Internet]. 2023 [cited 2023 Jun 30]. Available from: https://www.who.int/news/item/28-03-2023-sage-updates-covid-19-vaccination-guidance

